# Risk factors for severe COVID-19 in hospitalized children in Canada: A national prospective study from March 2020–May 2021

**DOI:** 10.1101/2022.04.06.22273409

**Authors:** Daniel S. Farrar, Olivier Drouin, Charlotte Moore Hepburn, Krista Baerg, Kevin Chan, Claude Cyr, Elizabeth J. Donner, Joanne E. Embree, Catherine Farrell, Sarah Forgie, Ryan Giroux, Kristopher T. Kang, Melanie King, Melanie Laffin Thibodeau, Julia Orkin, Naïm Ouldali, Jesse Papenburg, Catherine M. Pound, Victoria E. Price, Jean-Philippe Proulx-Gauthier, Rupeena Purewal, Christina Ricci, Manish Sadarangani, Marina I. Salvadori, Roseline Thibeault, Karina A. Top, Isabelle Viel-Thériault, Fatima Kakkar, Shaun K. Morris

## Abstract

**Background:** Children living with chronic comorbid conditions are at increased risk for severe COVID-19, though there is limited evidence regarding the risks associated with specific conditions and which children may benefit from targeted COVID-19 therapies. The objective of this study was to identify factors associated with severe disease among hospitalized children with COVID-19 in Canada.

**Methods:** We conducted a national prospective study on hospitalized children with microbiologically confirmed SARS-CoV-2 infection via the Canadian Paediatric Surveillance Program from April 2020–May 2021. Cases were reported voluntarily by a network of >2800 paediatricians. Hospitalizations were classified as COVID-19-related, incidental infection, or infection control/social admissions. Severe disease (among COVID-19-related hospitalizations only) was defined as disease requiring intensive care, ventilatory or hemodynamic support, select organ system complications, or death. Risk factors for severe disease were identified using multivariable Poisson regression, adjusting for age, sex, concomitant infections, and timing of hospitalization.

**Findings:** We identified 544 children hospitalized with SARS-CoV-2 infection, including 60·7% with COVID-19-related disease and 39·3% with incidental infection or infection control/social admissions. Among COVID-19-related hospitalizations (n=330), the median age was 1·9 years (IQR 0·1–13·3) and 43·0% had chronic comorbid conditions. Severe disease occurred in 29·7% of COVID-19-related hospitalizations (n=98/330), most frequently among children aged 2-4 years (48·7%) and 12-17 years (41·3%). Comorbid conditions associated with severe disease included technology dependence (adjusted risk ratio [aRR] 2·01, 95% confidence interval [CI] 1·37-2·95), neurologic conditions (e.g. epilepsy and select chromosomal/genetic conditions) (aRR 1·84, 95% CI 1·32-2·57), and pulmonary conditions (e.g. bronchopulmonary dysplasia and uncontrolled asthma) (aRR 1·63, 95% CI 1·12-2·39).

**Interpretation:** While severe outcomes were detected at all ages and among patients with and without comorbidities, neurologic and pulmonary conditions as well as technology dependence were associated with increased risk of severe COVID-19. These findings may help guide vaccination programs and prioritize targeted COVID-19 therapies for children.

**Funding:** Financial support for the CPSP was received from the Public Health Agency of Canada.

## BACKGROUND

Infection with severe acute respiratory syndrome coronavirus 2 (SARS-CoV-2) is typically mild or asymptomatic in children as compared to adults, however, severe outcomes including hospitalization and death due to coronavirus disease 2019 (COVID-19) have been reported.^1,2^ Chronic comorbid conditions are an important prognostic factor for disease progression, associated with odds of severe disease two to four times higher than children without these conditions.^3,4^ Evidence regarding risk associated with specific conditions remains limited, though diabetes, neurologic and respiratory conditions, and multiple comorbidities have emerged as conditions associated with high risk of severe COVID-19.^5–7^ Age-based estimates of risk have been mixed and are often subject to jurisdictional differences in admission thresholds, heterogeneous definitions of severity, and may be complicated by the inclusion of patients with multisystem inflammatory syndrome in children (MIS-C).^6,8–10^ Robust estimates of specific risk factors in children are needed to inform evidence-based decision-making by clinicians, policymakers, and families.

In Canada, more than 1·3 million laboratory-confirmed cases of SARS-CoV-2 infection across all ages were reported as of May 31, 2021, including successive waves of the original SARS-CoV-2 strain, Alpha (B.1.1.7) variant, and early cases of the Delta (B.1.617.2) variant.^11^ COVID-19 vaccination and booster programs have since been introduced, including stepwise approvals for children aged ≥16 years, 12-15 years, and 5-11 years.^12^ Select treatments have also been shown to decrease the morbidity and mortality burden of COVID-19 in adults, including corticosteroids and other immunomodulators as well as novel therapies (e.g. remdesivir, nirmatrelvir, and monoclonal antibodies). However, indications guiding the use of these agents in children are less clear.^13,14^ Age-specific baseline indicators of COVID-19 severity including hospitalization, intensive care unit (ICU) admission, and respiratory support requirements are therefore needed to a) evaluate severity trends amidst ongoing vaccination strategies, and b) guide the use of targeted COVID-19 therapies in children.

In this study, we present final findings from a nationally representative prospective surveillance study of hospitalized children with microbiologically confirmed SARS-CoV-2 infection in Canada, prior to emergence of the Omicron variant and the approval of SARS-CoV-2 vaccines for use in children. The primary objectives were to 1) identify specific comorbid conditions associated with severe COVID-19 among hospitalized Canadian children and 2) describe severity within specific paediatric age groups.

## METHODS

### Study design and procedures

The Canadian Paediatric Surveillance Program (CPSP) is a public health surveillance network which actively collects national, population-based data regarding rare childhood disorders, and is jointly operated by the Canadian Paediatric Society (CPS) and Public Health Agency of Canada (PHAC).^15^ This network includes >2,800 paediatricians and paediatric subspecialists across Canada who report cases to CPSP on a voluntary basis. The CPSP COVID-19 study launched on April 8, 2020 and asked all participating physicians to report any incident cases on a weekly basis using online case reporting via the Canadian Network for Public Health Intelligence. Cases of children <18 years of age and hospitalized with microbiologically-confirmed SARS-CoV-2 infection between the onset of the COVID-19 pandemic until May 31, 2021 were eligible to be reported. For this analysis, we excluded cases meeting criteria for MIS-C who tested positive by polymerase chain reaction (PCR) during their hospital stay. A preliminary analysis of cases occurring from March–December 2020 has previously been published.^16^ The protocol and case report form for the study can be found at https://cpsp.cps.ca/surveillance/study-etude/covid-19.

For all cases, participating physicians were asked to report on demographic characteristics, SARS-CoV-2 testing and exposures, chronic comorbid conditions, clinical features including the reason for hospitalization and presenting symptoms, and outcomes including level of care required and supports/treatments administered. Participants could consent to follow-up from the study team for clarification in case of incomplete or discrepant data elements, and consent was provided for 96% of reported cases. Real-time data management including record de-duplication, cleaning, and participant follow-up was conducted throughout the study period.

### Study definitions

The reason for hospitalization was physician-reported as either a) COVID-19-related (any child whose presentation was clinically consistent with symptomatic COVID-19 and who required admission because of those symptoms); or b) not related to COVID-19 (any child admitted for another condition in whom SARS-CoV-2 was detected upon routine screening, or a child admitted for isolation/infection control or social reasons, and whose SARS-CoV-2 symptoms would not otherwise have warranted hospitalization). For each case, at least two study investigators (OD, CMH, FK, or SKM) reviewed all available clinical details to ensure consistency in the physician-reported categories, with any discrepancies in interpretation resolved via consensus discussion. Among patients hospitalized for COVID-19, disease severity was then assigned using an algorithm adapted from the World Health Organization^17^ and cases were categorized as either mild disease (symptomatic COVID-19 not requiring supplemental oxygen or targeted therapy); moderate disease (requiring oxygen above baseline home needs or targeted therapy such as remdesivir or corticosteroids); or severe disease (admitted to intensive care, requiring non-invasive or mechanical ventilation or vasopressors, experiencing respiratory, neurologic, or cardiac organ complications, or any recorded death); (Appendix 1). Disease severity was then analyzed as a binary outcome whereby mild and moderate disease were aggregated as ‘non-severe’ disease.

Chronic comorbid conditions were first analyzed as specific conditions or subcategories and included: chronic renal disease, congenital heart disease, diabetes mellitus, gastrointestinal disease, hematologic disease, immunosuppression, malignancies, metabolic disease, neurologic or neurodevelopmental disorders, obesity, psychiatric disorders, pulmonary diseases, technology dependence, and transplant recipients. Immunosuppression was defined as any immunosuppressing medications (including active chemotherapy), primary or secondary immunodeficiency, or chronic rheumatologic or autoimmune disorders. Technology dependence included presence of tracheostomy and/or requirements for home oxygen, parenteral nutrition, or dialysis. Obesity was categorized using body mass index-for-age Z-scores (BMIZ) according to the WHO Child Growth Standards.^18^

To assess overall medical complexity, we assigned patients to one of three mutually exclusive categories using a definition informed by prior literature.^19,20^ For this indicator, ‘complex chronic disease’ includes a) patients with two or more comorbid conditions occurring in multiple body systems, or b) patients with a single comorbid condition which impacts multiple body systems, has technology dependence requirements, or is associated with a shorter than average lifespan; ‘non-complex chronic disease’ includes all other patients with comorbid conditions; and ‘none/unknown’ includes patients with no known or confirmed comorbid conditions.

Child age was categorized as <6 months, 6-23 months, 2-4 years, 5-11 years, and 12-17 years. Timing of hospitalization was categorized as first wave (March–August 2020), second wave (September 2020–February 2021), or third wave (March–May 2021). Summaries of all radiologic findings were reviewed by investigators (as above) and categorized as either abnormal, abnormal but not related to SARS-CoV-2, non-specific, or normal.

### Statistical analysis

Demographic and clinical characteristics were summarized using frequencies, percentages, medians, and interquartile ranges (IQR). Due to CPSP privacy policies, frequencies between one and four were masked and reported as ‘<5’ while some larger frequencies were presented as ranges to prevent back-calculation. Subgroup comparisons were analyzed using χ^2^ tests, Fisher’s exact tests, and Wilcoxon rank-sum tests, as appropriate. Analysis of risk factors and severity outcomes was restricted to patients hospitalized with COVID-19-related disease. Multivariable Poisson regression with robust standard errors was used to identify risk factors for severe COVID-19, reported using adjusted risk ratios (aRR). The primary adjusted model, defined *a priori*, included age, sex, concomitant infections, timing of hospitalization, and chronic conditions (categorized as none, non-complex, or complex). Child age was entered as a continuous variable using a restricted cubic spline with four knots at evenly-spaced percentiles.

To assess the role of specific conditions, the chronic comorbid condition variable was substituted with each individual condition or subcategory and analyzed as described above. A separate model was conducted among children <1 year only to assess very young age (i.e. <1 month versus 1-11 months) and premature births (vs. term births), adjusting only for those variables due to a smaller available sample size. Minimum population-based incidence proportions were calculated by dividing the number of age-specific hospitalizations and severe COVID-19 cases by 2020 midyear population denominators from Statistics Canada.^21^ Confidence intervals (CI) were computed by assuming a Poisson distribution. Data analysis was conducted in Stata version 17·0, using a statistical significance threshold of α=0·05.^22^

### Ethical approval

The CPSP operates under the authority derived from Section 4 of the Department of Health Act and Section 3 of the PHAC act. Approval was obtained from Research Ethics Boards at Health Canada-PHAC (REB #2020-002P), the Hospital for Sick Children (REB #1000070001), the Centre Hospitalier Universitaire Sainte-Justine (IRB #MP-21-2021-2901), and at individual sites as required by local policies.

## RESULTS

During the fifteen-month study period, 544 children hospitalized with SARS-CoV-2 infection met this study’s inclusion criteria (Figure S1). Among these cases, 330 (60·7%) were hospitalized with COVID-19-related disease, while the remainder were incidental cases admitted for unrelated care (n=201, 36·9%) or admitted for infection control or social purposes (n=13, 2·4%). Overall, 15·3% of cases were hospitalized during the first pandemic wave (peaking in April 2020), 50·0% during the second wave (peaking in January 2021), and 34·7% during the third wave (peaking in April 2021; Figure S2). Hospitalizations were reported from all regions across Canada, most commonly from the most populous provinces of Ontario (n=229; 42·1%) and Quebec (n=194; 35·7%).

Of the 330 COVID-19-related hospitalizations, 70·3% (n=232) met criteria for non-severe disease while 29·7% (n=98) met criteria for severe disease (Table 1). The median age at admission was 1·9 years (IQR 0·1–13·3) and was lower among patients with non-severe COVID-19 (0·8 years, IQR 0·1–9·7) than those with severe COVID-19 (6·5 years, IQR 1·5–14·8; p<0·001). Accounting for underlying population size, study period incidence proportions for COVID-19 hospitalization and severe COVID-19 were highest among children <1 year of age (37·9 hospitalizations and 5·4 severe cases per 100,000 population) and lowest among children 5-11 years of age (1·0 hospitalizations and 0·4 severe cases per 100,000 population; Table 2). Concomitant infections were reported among 8·2% of cases (n=27), including most commonly urinary tract infections (n=10). Among 140 patients <1 year old, 12·9% (n=18) were born at <37 weeks gestation.

**Table 1.**
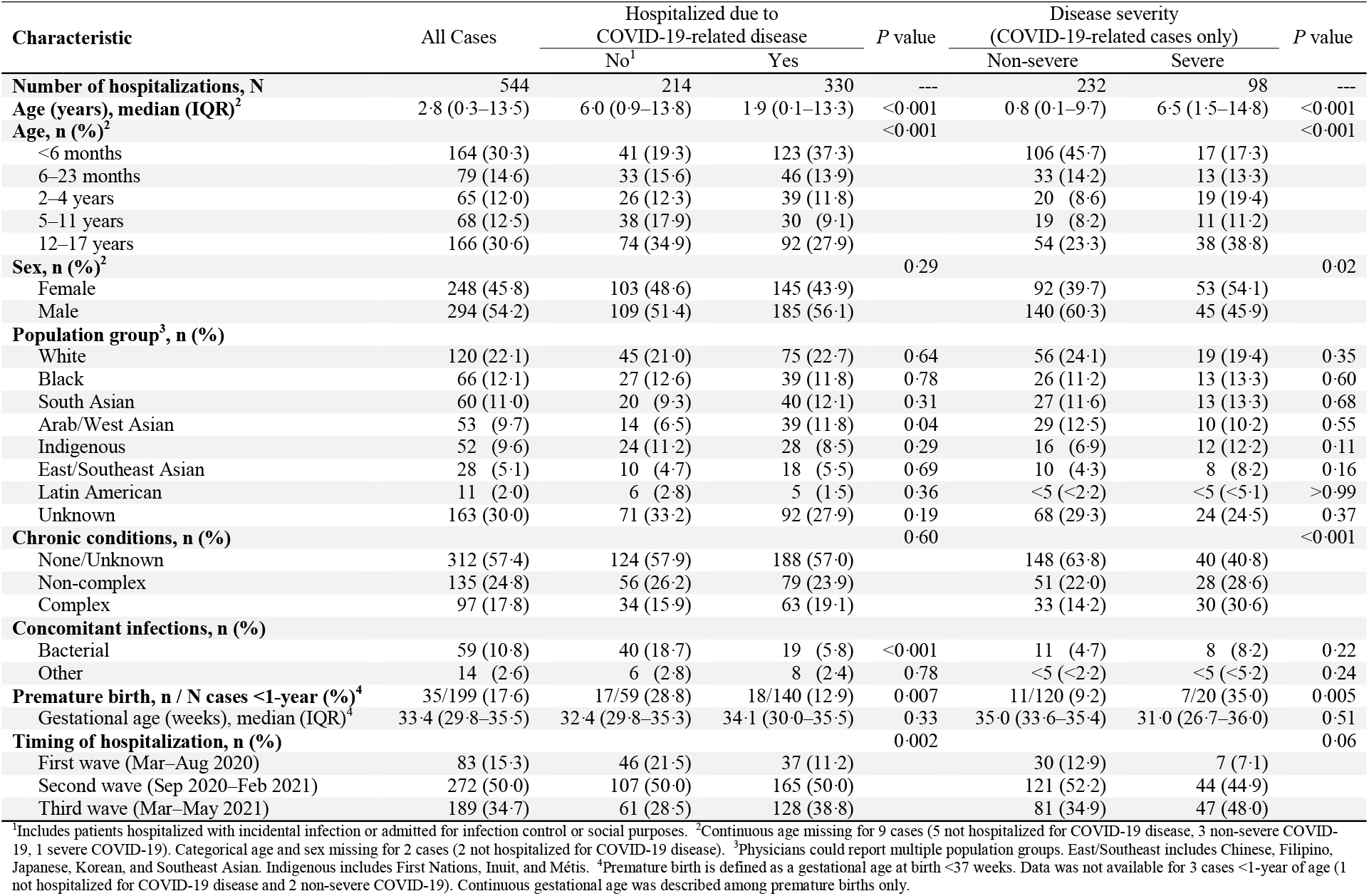
Characteristics of children hospitalized with SARS-CoV-2 infection in Canada until May 31, 2021.

| Characteristic | All Cases | Hospitalized due to<br>COVID-19-related disease |  | *P* value | Disease severity<br>(COVID-19-related cases only) |  | *P* value |
| --- | --- | --- | --- | --- | --- | --- | --- |
|  |  | No<sup>1</sup> | Yes |  | Non-severe | Severe |  |
| **Number of hospitalizations, N** | 544 | 214 | 330 | --- | 232 | 98 | --- |
| **Age (years), median (IQR)<sup>2</sup>** | 2.8 (0.3–13.5) | 6.0 (0.9–13.8) | 1.9 (0.1–13.3) | <0.001 | 0.8 (0.1–9.7) | 6.5 (1.5–14.8) | <0.001 |
| **Age, n (%)<sup>2</sup>** |  |  |  | <0.001 |  |  | <0.001 |
| <6 months | 164 (30.3) | 41 (19.3) | 123 (37.3) |  | 106 (45.7) | 17 (17.3) |  |
| 6–23 months | 79 (14.6) | 33 (15.6) | 46 (13.9) |  | 33 (14.2) | 13 (13.3) |  |
| 2–4 years | 65 (12.0) | 26 (12.3) | 39 (11.8) |  | 20 (8.6) | 19 (19.4) |  |
| 5–11 years | 68 (12.5) | 38 (17.9) | 30 (9.1) |  | 19 (8.2) | 11 (11.2) |  |
| 12–17 years | 166 (30.6) | 74 (34.9) | 92 (27.9) |  | 54 (23.3) | 38 (38.8) |  |
| **Sex, n (%)<sup>2</sup>** |  |  |  | 0.29 |  |  | 0.02 |
| Female | 248 (45.8) | 103 (48.6) | 145 (43.9) |  | 92 (39.7) | 53 (54.1) |  |
| Male | 294 (54.2) | 109 (51.4) | 185 (56.1) |  | 140 (60.3) | 45 (45.9) |  |
| **Population group<sup>3</sup>, n (%)** |  |  |  |  |  |  |  |
| White | 120 (22.1) | 45 (21.0) | 75 (22.7) | 0.64 | 56 (24.1) | 19 (19.4) | 0.35 |
| Black | 66 (12.1) | 27 (12.6) | 39 (11.8) | 0.78 | 26 (11.2) | 13 (13.3) | 0.60 |
| South Asian | 60 (11.0) | 20 (9.3) | 40 (12.1) | 0.31 | 27 (11.6) | 13 (13.3) | 0.68 |
| Arab/West Asian | 53 (9.7) | 14 (6.5) | 39 (11.8) | 0.04 | 29 (12.5) | 10 (10.2) | 0.55 |
| Indigenous | 52 (9.6) | 24 (11.2) | 28 (8.5) | 0.29 | 16 (6.9) | 12 (12.2) | 0.11 |
| East/Southeast Asian | 28 (5.1) | 10 (4.7) | 18 (5.5) | 0.69 | 10 (4.3) | 8 (8.2) | 0.16 |
| Latin American | 11 (2.0) | 6 (2.8) | 5 (1.5) | 0.36 | <5 (<2.2) | <5 (<5.1) | >0.99 |
| Unknown | 163 (30.0) | 71 (33.2) | 92 (27.9) | 0.19 | 68 (29.3) | 24 (24.5) | 0.37 |
| **Chronic conditions, n (%)** |  |  |  | 0.60 |  |  | <0.001 |
| None/Unknown | 312 (57.4) | 124 (57.9) | 188 (57.0) |  | 148 (63.8) | 40 (40.8) |  |
| Non-complex | 135 (24.8) | 56 (26.2) | 79 (23.9) |  | 51 (22.0) | 28 (28.6) |  |
| Complex | 97 (17.8) | 34 (15.9) | 63 (19.1) |  | 33 (14.2) | 30 (30.6) |  |
| **Concomitant infections, n (%)** |  |  |  |  |  |  |  |
| Bacterial | 59 (10.8) | 40 (18.7) | 19 (5.8) | <0.001 | 11 (4.7) | 8 (8.2) | 0.22 |
| Other | 14 (2.6) | 6 (2.8) | 8 (2.4) | 0.78 | <5 (<2.2) | <5 (<5.2) | 0.24 |
| **Premature birth, n / N cases <1-year (%)<sup>4</sup>** | 35/199 (17.6) | 17/59 (28.8) | 18/140 (12.9) | 0.007 | 11/120 (9.2) | 7/20 (35.0) | 0.005 |
| Gestational age (weeks), median (IQR)<sup>4</sup> | 33.4 (29.8–35.5) | 32.4 (29.8–35.3) | 34.1 (30.0–35.5) | 0.33 | 35.0 (33.6–35.4) | 31.0 (26.7–36.0) | 0.51 |
| **Timing of hospitalization, n (%)** |  |  |  | 0.002 |  |  | 0.06 |
| First wave (Mar–Aug 2020) | 83 (15.3) | 46 (21.5) | 37 (11.2) |  | 30 (12.9) | 7 (7.1) |  |
| Second wave (Sep 2020–Feb 2021) | 272 (50.0) | 107 (50.0) | 165 (50.0) |  | 121 (52.2) | 44 (44.9) |  |
| Third wave (Mar–May 2021) | 189 (34.7) | 61 (28.5) | 128 (38.8) |  | 81 (34.9) | 47 (48.0) |  |

<sup>1</sup>Includes patients hospitalized with incidental infection or admitted for infection control or social purposes. <sup>2</sup>Continuous age missing for 9 cases (5 not hospitalized for COVID-19 disease, 3 non-severe COVID-19, 1 severe COVID-19). Categorical age and sex missing for 2 cases (2 not hospitalized for COVID-19 disease). <sup>3</sup>Physicians could report multiple population groups. East/Southeast includes Chinese, Filipino, Japanese, Korean, and Southeast Asian. Indigenous includes First Nations, Inuit, and Métis. <sup>4</sup>Premature birth is defined as a gestational age at birth <37 weeks. Data was not available for 3 cases <1-year of age (1 not hospitalized for COVID-19 disease and 2 non-severe COVID-19). Continuous gestational age was described among premature births only.

**Table 2.** Minimum population-based incidence of SARS-CoV-2 and COVID-19-related outcomes from March 2020–May 2021 in Canada.

| Category | Minimum incidence, Mar 2020–May 2021<br>(per 100,000 population; 95% CI)<sup>1</sup> |
| --- | --- |
| **All SARS-CoV-2 hospitalizations** |  |
| <1 year | 53.9 (46.7–61.9) |
| 1–4 years | 7.0 (5.8–8.5) |
| 5–11 years | 2.4 (1.8–3.0) |
| 12–17 years | 6.8 (5.8–7.9) |
| **COVID-19-related hospitalizations** |  |
| <1 year | 37.9 (31.9–44.7) |
| 1–4 years | 4.4 (3.4–5.6) |
| 5–11 years | 1.0 (0.7–1.5) |
| 12–17 years | 3.8 (3.0–4.6) |
| **Severe COVID-19** |  |
| <1 year | 5.4 (3.3–8.4) |
| 1–4 years | 1.9 (1.3–2.7) |
| 5–11 years | 0.4 (0.2–0.7) |
| 12–17 years | 1.6 (1.1–2.1) |

<sup>1</sup>Minimum incidence was calculated using age band-specific midyear population denominators for 2020, retrieved from Statistics Canada. Confidence intervals (CI) were calculated using the Poisson distribution.

Children living with comorbid conditions comprised 43·0% of COVID-19 hospitalizations (n=142), including 23·9% (n=79) classified as non-complex conditions and 19.1% (n=63) classified as complex conditions (Table 1). Neurologic and neurodevelopmental disorders were most common (n=46, 13·9%) and included epilepsy (n=20), chronic encephalopathies (n=19 including 8 with cerebral palsy), and chromosomal/genetic disorders (n=9 including <5 with trisomy 21, Table 3). Forty-four patients (13·3%) were obese, all aged ≥5 years, with a median BMIZ of 2·9 (among 22 obese patients with reported height and weight data, IQR 2·3–3·3). Pulmonary conditions were reported in 34 patients (10·3%), including asthma most commonly (n=16; 8 using daily controller medications and 8 not using daily controller medications). There were few reported with immunosuppression (n=19, 5·8%; including 14 with immunosuppressing medications), hematologic disorders (n=17, 5·2%; including 14 with sickle cell disease), congenital heart disease (n=14, 4·2%), gastrointestinal/liver disease (n=10, 3·0%), diabetes (n=8 including 7 who were insulin dependent), and malignancies (n=8, 2·4%; including 6 with leukemia). Finally, 16 patients (4·8%) had pre-existing technology dependence requirements including 12 requiring parenteral nutrition and seven with either tracheostomy or home oxygen requirements. Compared to patients hospitalized for other reasons, children hospitalized for COVID-19 were more likely to be obese (13·3% vs. 3·7%, p<0·001) and have pulmonary conditions (10·3% vs. 3·3%, p=0·002) but less likely to have psychiatric disorders (<1·5% vs. 4·2%, p=0·001; Table S1).

**Table 3.** Chronic comorbid conditions among COVID-19-related hospitalizations.

| Chronic comorbid conditions, n (%) | All COVID-19 related cases | Disease severity |  | P value |
| --- | --- | --- | --- | --- |
|  |  | Non-severe | Severe |  |
| **Number of hospitalizations, N** | 330 | 232 | 98 | --- |
| **Neurologic/neurodevelopmental disorder, any<sup>1</sup>** | 46 (13.9) | 18 (7.8) | 28 (28.6) | <0.001 |
| Epilepsy | 20 (6.1) | 5 (2.2) | 15 (15.3) | <0.001 |
| Chronic encephalopathy (e.g. cerebral palsy) | 19 (5.8) | 7 (3.0) | 12 (12.2) | 0.001 |
| Chromosomal/genetic disorders | 9 (2.7) | <5 (<2.2) | 5–8 (5.1–8.2) | 0.004 |
| Neurologic, NOS | 12 (3.6) | 7 (3.0) | 5 (5.1) | 0.35 |
| **Obesity<sup>2</sup>** | 44 (13.3) | 21 (9.1) | 23 (23.5) | <0.001 |
| **Pulmonary, any<sup>1</sup>** | 34 (10.3) | 16 (6.9) | 18 (18.4) | 0.002 |
| Asthma | 16 (4.8) | 8 (3.4) | 8 (8.2) | 0.09 |
| Bronchopulmonary dysplasia | 9 (2.7) | <5 (<2.2) | 5–8 (5.1–8.2) | 0.02 |
| Pulmonary, NOS | 10 (3.0) | 5 (2.2) | 5 (5.1) | 0.17 |
| **Immunosuppression, any<sup>1,3</sup>** | 19 (5.8) | 15–18 (6.5–7.8) | <5 (<5.1) | 0.17 |
| Immunocompromising medications | 14 (4.2) | 10–13 (4.3–5.6) | <5 (<5.1) | 0.77 |
| **Hematologic disease, any<sup>1</sup>** | 17 (5.2) | 13–16 (5.6–6.9) | <5 (<5.1) | 0.10 |
| Sickle cell disease | 14 (4.2) | 10–13 (4.3–5.6) | <5 (<5.1) | 0.07 |
| **Technology dependence, any<sup>1,4</sup>** | 16 (4.8) | 5 (2.2) | 11 (11.2) | 0.001 |
| Parenteral nutrition | 12 (3.6) | 5 (2.2) | 7 (7.1) | 0.047 |
| Respiratory technology dependence | 7 (2.1) | 0 (0.0) | 7 (7.1) | <0.001 |
| **Congenital heart disease** | 14 (4.2) | 7 (3.0) | 7 (7.1) | 0.13 |
| **Gastrointestinal/liver disease** | 10 (3.0) | 6–9 (2.6–3.9) | <5 (<5.1) | >0.99 |
| **Chronic renal disease** | 9 (2.7) | 5–8 (2.2–3.4) | <5 (<5.1) | 0.73 |
| **Diabetes mellitus** | 8 (2.4) | <5 (<2.2) | <5 (<5.1) | 0.24 |
| **Malignancy** | 8 (2.4) | 5–7 (2.2–3.0) | <5 (<5.1) | >0.99 |
| **Metabolic disease** | 8 (2.4) | 5–7 (2.2–3.0) | <5 (<5.1) | 0.70 |
| **Bone diseases** | <5 (<1.5) | DNS | DNS | --- |
| **Other perinatal conditions** | <5 (<1.5) | DNS | DNS | --- |
| **Psychiatric disorder** | <5 (<1.5) | DNS | DNS | --- |
| **Transplant recipient** | <5 (<1.5) | DNS | DNS | --- |

DNS = Data not shown due to <5 frequencies across multiple subgroups; NOS = Not otherwise specified.
<sup>1</sup>Multiple specific conditions could be reported (e.g. two or more neurologic conditions) and therefore may not sum to category total.
<sup>2</sup>Obesity was physician-reported and could not be confirmed for children missing height and weight data (n=22).
<sup>3</sup>Defined as immunocompromising medications, primary or secondary immunodeficiency otherwise specified, or chronic rheumatologic or autoimmune disorder.
<sup>4</sup>Defined as parenteral nutrition, respiratory technology requirements (i.e. home oxygen or tracheostomy), or dialysis.

Among COVID-19-related hospitalizations, 60 (18·2%) children were admitted to the ICU for a median duration of four days (IQR 2–7, Table 4). Including patients admitted to the ward and ICU, nearly one-third (n=108) required respiratory or hemodynamic support, including 19·1% (n=63) who required support greater than low-flow oxygen. Mechanical ventilation was required for 7·6% of children (n=25), while few (n=8, 2·4%) required vasopressors and none required extracorporeal membrane oxygenation. Among children requiring mechanical ventilation, 11/25 had no known chronic comorbid conditions (including 5/10 children aged 2-4 years). Five children died due to complications of acute COVID-19, at a mean age of 8·1 years (standard deviation 7·3 years). Children aged 2-4 years experienced the highest proportion of severe disease (48·7%, n=19/39), followed by those aged 12-17 years (41·3%, n=38/92), 5-11 years (36·7%, n=11/30), 6-23 months (28·3%, n=13/46), and finally <6 months (13·8%, n=17/123). Children aged 2-4 years were more often admitted to ICU (33·3%, n=13/39) while children aged 12-17 years more often required any respiratory or hemodynamic support (51·1%, n=47/92). The proportion of children with chronic comorbid conditions by age group is described in Table S2, while severity and treatment outcomes by pandemic wave is described in Table S3.

**Table 4.** Severity and treatment outcomes of children hospitalized with COVID-19 in Canada, by age group.

| Characteristics | All Cases | Child age |  |  |  |  |
| --- | --- | --- | --- | --- | --- | --- |
|  |  | <6 months | 6–23 months | 2–4 years | 5–11 years | 12–17 years |
| **COVID-19-related hospitalizations, N** | 330 | 123 | 46 | 39 | 30 | 92 |
| **COVID-19 severity, n (%)** |  |  |  |  |  |  |
| Mild/moderate illness | 232 (70.3) | 106 (86.2) | 33 (71.7) | 20 (51.3) | 19 (63.3) | 54 (58.7) |
| Severe illness | 98 (29.7) | 17 (13.8) | 13 (28.3) | 19 (48.7) | 11 (36.7) | 38 (41.3) |
| **Admitted to ICU, n (%)** | 60 (18.2) | 12 (9.8) | 7 (15.2) | 13 (33.3) | 7 (23.3) | 21 (22.8) |
| Length of ICU stay, median (IQR) | 4 (2–7) | 3 (3–10) | 4 (3–6) | 2 (2–4) | 2 (1–9) | 6 (3–8) |
| **Respiratory/hemodynamic support required, n (%)<sup>1</sup>** | 108 (32.7) | 20 (16.3) | 15 (32.6) | 17 (43.6) | 9 (30.0) | 47 (51.1) |
| Low-flow oxygen | 58 (17.6) | 13 (10.6) | 8 (17.4) | 6 (15.4) | 6 (20.0) | 25 (27.2) |
| High-flow nasal cannula | 33 (10.0) | 5 (4.1) | <5 (<10.9) | 5 (12.8) | <5 (<16.7) | 15 (16.3) |
| Non-invasive ventilation (e.g. CPAP or BiPAP) | 14 (4.2) | <5 (<4.1) | <5 (<10.9) | <5 (<12.8) | <5 (<16.7) | 5 (5.4) |
| Conventional mechanical ventilation | 25 (7.6) | <5 (<4.1) | <5 (<10.9) | 10 (25.6) | 0 (0.0) | 8 (8.7) |
| Vasopressors | 8 (2.4) | 0 (0.0) | <5 (<10.9) | <5 (<12.8) | 0 (0.0) | <5 (<5.4) |
| **Clinical presentation, n (%)** |  |  |  |  |  |  |
| Fever | 241 (73.0) | 90 (73.2) | 40 (87.0) | 31 (79.5) | 20 (66.7) | 60 (65.2) |
| Upper respiratory tract infection | 202 (61.2) | 71 (57.7) | 29 (63.0) | 19 (48.7) | 15 (50.0) | 68 (73.9) |
| Gastrointestinal | 125 (37.9) | 37 (30.1) | 16 (34.8) | 14 (35.9) | 14 (46.7) | 44 (47.8) |
| Hematologic disorder | 90 (27.3) | 26 (21.1) | 11 (23.9) | 10 (25.6) | 10 (33.3) | 33 (35.9) |
| Pneumonia | 84 (25.5) | 9 (7.3) | 13 (28.3) | 9 (23.1) | 10 (33.3) | 43 (46.7) |
| Coagulation dysfunction | 22 (6.7) | <5 (<4.1) | <5 (<10.9) | <5 (<12.8) | <5 (<16.7) | 13 (14.1) |
| Seizure(s) | 22 (6.7) | <5 (<4.1) | <5 (<10.9) | 9 (23.1) | <5 (<16.7) | 6 (6.5) |
| Acute respiratory distress syndrome | 18 (5.5) | <5 (<4.1) | <5 (<10.9) | <5 (<12.8) | 0 (0.0) | 12 (13.0) |
| Bronchiolitis | 18 (5.5) | 11 (8.9) | 7 (15.2) | 0 (0.0) | 0 (0.0) | 0 (0.0) |
| Hypotension | 15 (4.6) | 0 (0.0) | <5 (<10.9) | <5 (<12.8) | <5 (<16.7) | 6 (6.5) |
| Hepatitis | 14 (4.2) | <5 (<4.1) | <5 (<10.9) | <5 (<12.8) | <5 (<16.7) | 7 (7.6) |
| Renal dysfunction | 14 (4.2) | 0 (0.0) | <5 (<10.9) | <5 (<12.8) | <5 (<16.7) | 7 (7.6) |
| Acute cardiac dysfunction | 6 (1.8) | DNS | DNS | DNS | DNS | DNS |
| Cytokine storm/MAS | 5 (1.5) | DNS | DNS | DNS | DNS | DNS |
| Coma | <5 (<1.5) | DNS | DNS | DNS | DNS | DNS |
| Encephalopathy | <5 (<1.5) | DNS | DNS | DNS | DNS | DNS |
| **Abnormal CXR, n / cases with imaging (%)** | 100/224 (44.6) | 13/74 (17.6) | 14/30 (46.7) | 14/28 (50.0) | 9/21 (42.9) | 48/71 (67.6) |
| **COVID-19-related therapies, n (%)** |  |  |  |  |  |  |
| Steroids | 88 (26.7) | 5 (4.1) | 14 (30.4) | 11 (28.2) | 11 (36.7) | 47 (51.1) |
| Anticoagulation | 37 (11.2) | 0 (0.0) | <5 (<10.9) | <5 (<12.8) | <5 (<16.7) | 28 (30.4) |
| Remdesivir | 32 (9.7) | <5 (<4.1) | <5 (<10.9) | 6 (15.4) | <5 (<16.7) | 18 (19.6) |
| Biologics | 7 (2.1) | DNS | DNS | DNS | DNS | DNS |
| Immunoglobulin | 5 (1.5) | DNS | DNS | DNS | DNS | DNS |
| **Child died, n (%)** | 5 (1.5) | DNS | DNS | DNS | DNS | DNS |

BiPAP=Bilevel positive airway pressure; CPAP=Continuous positive airway pressure; CXR = Chest x-ray; DNS = Data not shown due to <5 frequencies across multiple subgroups;
ICU=Intensive care unit; IQR=Interquartile range; MAS=Macrophage activating syndrome. <sup>1</sup>Multiple supports could be reported and therefore specific supports do not sum to any supports.

There was a significant nonlinear relationship between age and severe COVID-19 (Figure 1), whereby children <1 year had significantly lower risk of severe disease while risk was highest for children aged 2-5 and 16-<18 years. In a separate model among children <1 year only, children aged <1 month had 2·55 times higher risk of severe disease (95% CI 1·15–5·64) than those aged 1-11 months. Several comorbid conditions were significantly associated with severe disease including any pulmonary condition (vs. none, aRR 1·63, 95% CI 1·12–2·39), any neurologic condition (vs. none, aRR 1·84, 95% CI 1·32–2·57), and any technology dependence requirements (vs. none, aRR 2·01, 95% CI 1·37–2·95; Figure 2). Bronchopulmonary dysplasia and epilepsy were the strongest risk factors for severe disease (aRR 2·39 [1·37–4·18] and 2·08 [1·44–2·99], respectively). Associations between asthma and COVID-19 severity were modified by the use of controller medications, as those with controlled asthma were not at higher risk of severe disease (aRR 0·33, 95% CI 0·05–2·20) while those with uncontrolled asthma were (aRR 2·24, 95% CI 1·54–3·27). Neurologic and pulmonary conditions as well as technology dependence requirements were often clustered together in the same patients with severe COVID-19 (Figure S3). Among children with obesity, only those with known BMIZ >3 had higher risk of severe disease (aRR 1·90, 95% CI 1·10–3·28). Immunosuppression (aRR 0·43, 95% CI 0·16– 1·16) and malignancies (aRR 0·65, 95% CI 0·22–1·90) were not associated with higher risk of severe COVID-19. Among patients <1 year old, prematurity increased the risk of severe disease by 3·47 times that of term-born children (95% CI 1·69–7·09). Notably, when analyzed as a single variable, non-complex conditions (aRR 1·10, 95% CI 0·71–1·69) and complex conditions (aRR 1·35, 95% CI 0·89–2·04) were not associated with higher risk of severe COVID-19 when compared to children with no known comorbid conditions.

**Figure 1.**
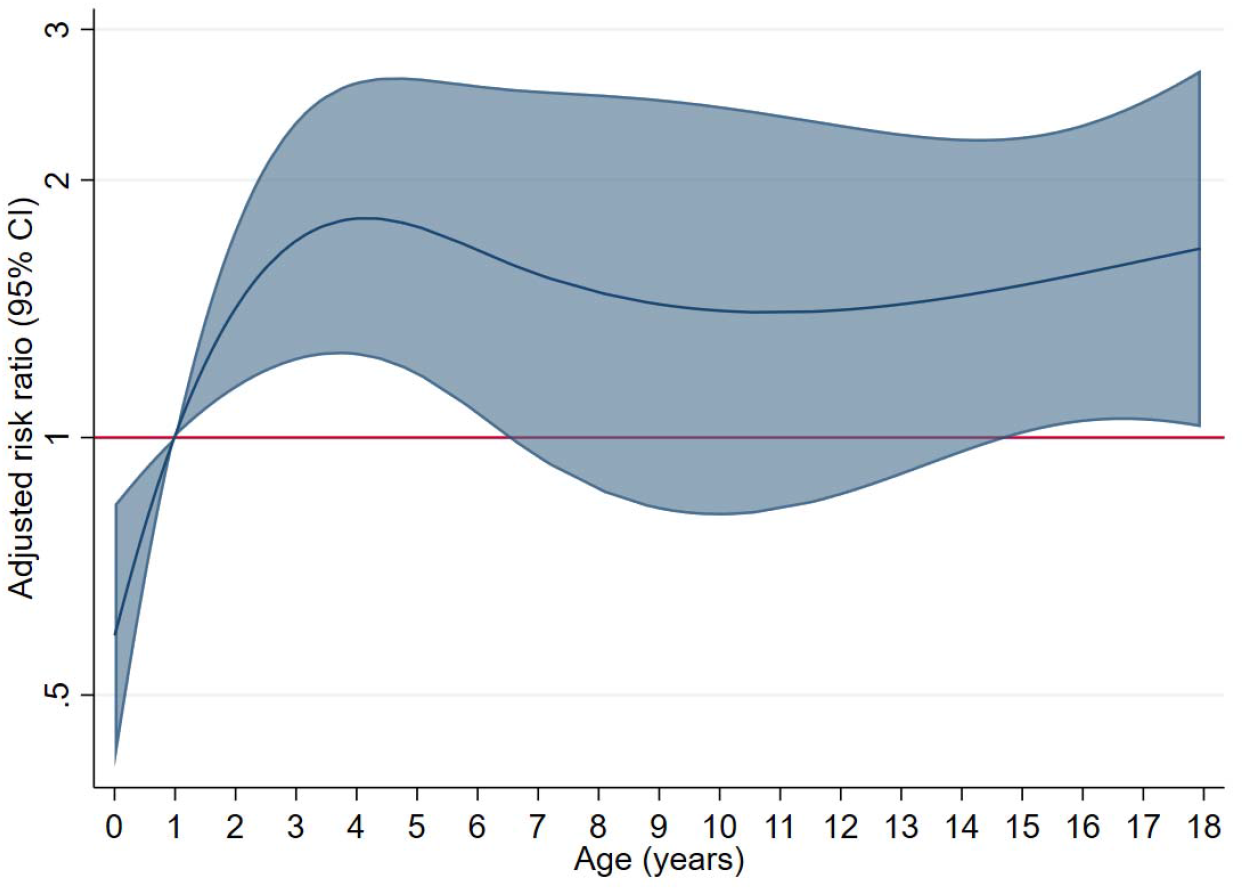
Risk ratios for severe COVID-19 by continuous child age. Age was analyzed using Poisson regression with robust standard errors, where age was entered into the model as a restricted cubic spline with four knots. Predicted probabilities of severe disease were then exponentiated to visualize continuous risk ratios. The analysis adjusted for sex, comorbid conditions (categorized as none, non-complex, or complex), concomitant infections (any vs. none), and timing of hospitalization (first, second, or third wave).

**Figure 2.**
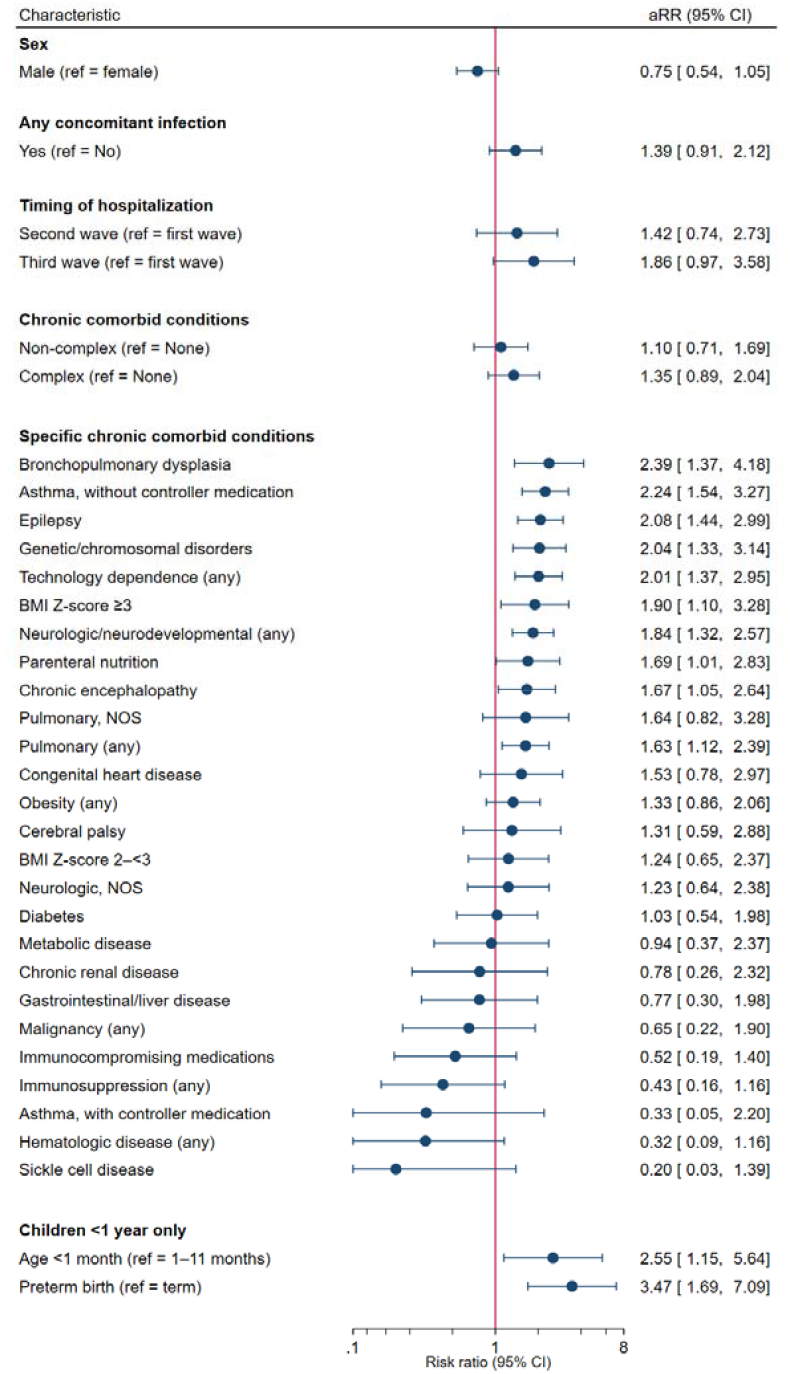
Risk ratios for severe COVID-19 by sex, concomitant infections, timing of hospitalization, and chronic conditions. Adjusted risk ratios (aRR) were calculated using Poisson regression with robust standard errors. The x-axis depicts risk ratios by factors of two (i.e. 2–4–6–8 and 1/2–1/4–1/6–1/8–1/10). The primary model included continuous age (as analyzed in Figure 1), sex, concomitant infections, timing of hospitalization, and chronic condition category (i.e. none/unknown, non-complex, complex). Separate models were then run for each specific chronic condition, by substituting the overall chronic condition category with only the condition of interest. Finally, age <1 month and prematurity status were assessed in a separate model containing only children <1 year old (n=140, including 20 severe cases), and did not adjust for additional variables due to a smaller available sample size.

## DISCUSSION

In this national prospective surveillance study, we described risk factors for disease severity and outcomes among 544 children hospitalized with SARS-CoV-2 infection (and without MIS-C) across Canada prior to emergence of the Omicron variant and the approval of paediatric COVID-19 vaccines. Overall, the CPSP voluntary reporting system captured 58% of children <18 years of age hospitalized with SARS-CoV-2 infection in Canada from March 2020–May 2021 (unpublished data; source: case information received by PHAC from provinces and territories).

While severe outcomes were rare in absolute terms, among COVID-19-related hospitalizations, one in three patients required respiratory or hemodynamic support (including one in five requiring greater than low-flow oxygen) while one in five required ICU admission, and five patients reported to the study died from their infection. Severe outcomes were identified across all age groups and among patients both with and without comorbid conditions. We estimated a lower proportion of severe COVID-19 than interim CPSP analyses (29·7% here vs. 50·0% in our prior report^16^). This was in part due to a change in definition from initial criteria adapted from Dong et al^23^ to modified WHO^17^ severity criteria, as application of the Dong criteria included subjective reports of organ involvement such as non-specific respiratory distress, coagulopathy, and minimal supplemental oxygen requirements (Table S4). Our results are similar to a US study of hospitalized patients with a primary diagnosis of COVID-19 (20·7% with severe disease), but the proportion with severe disease remained higher than most studies which included incidental cases in their study denominators (e.g. 4·1–15·4%).^24–26^ These differences may also be attributable to heterogeneous definitions of COVID-19 severity.

Child age was an important predictor of severe disease. After multivariable adjustment, hospitalized infants (i.e. children <1 year of age) had significantly lower risk of severe disease compared to older children, yet population-based incidence proportions of severe COVID-19 for this age group were between 3–18 times higher than older age groups. Lower in-hospital risk of severe disease is likely a result of lower thresholds for admission for febrile infants, in particular those <90 days old.^27^ Unlike most prior literature^8,10^, children aged 2-4 years in Canada exhibited the highest risk of severe disease, including 33% admitted to ICU and 26% requiring ventilation. These findings may inform vaccination strategies and use of COVID-19 therapies in this age group.

Chronic comorbid conditions associated with the neurologic or pulmonary systems had increased risk of severe disease roughly 1·5–2·5 times that of all other children. Epilepsy is now a well-established risk factor for severe disease amongst children, and may be caused by increased risk of seizures during infection as well as associated respiratory complications such as aspiration, and warrants dedicated study of specific epileptic subgroups.^5,28^ Technology dependence, most notably patients requiring home oxygen, tracheostomy, or gastrostomy tubes were at particularly high risk for severe COVID-19, consistent with past studies of other childhood respiratory viral infections such as influenza and respiratory syncytial virus.^29,30^ Notably, complex conditions as a single category were not significantly associated with severe COVID-19 risk, suggesting greater focus should be paid on specific conditions rather than broad categories as is often the case in risk stratification guidelines.^31,32^

Previous studies have shown mixed evidence of asthma as a risk factor for severe COVID-19.^5,7,10,24^ In this study, we demonstrate that controller medications may act as an effect modifier, whereby a lack of daily controller medications significantly increased the risk of severe disease. This finding suggests that asthma controllers may be protective against respiratory complications, and children lacking these medications may experience reduced access to care or be less likely to be actively followed in a clinical setting. This is supported by a recent population-based study that showed a marked increase in risk of SARS-CoV-2 hospitalization among school-aged children with poorly-controlled asthma.^33^

Unlike previous studies, diabetes was not associated with an increased risk of severe COVID-19.^5,10,26^ In this study, ten patients with diabetes (median age 12·6 years, IQR 4·6–14·7) were admitted for reasons other than COVID-19 including seven with diabetic ketoacidosis and it remains possible that SARS-CoV-2 infection precipitated or exacerbated these presentations.^34^ Confirming results from our interim analyses, children with suppressed immune systems including those with leukemia and other malignancies, sickle cell disease, and other immunocompromising conditions were not at higher risk of severe disease, consistent with case series of these specific disease groups.^35,36^

This study has several limitations. First, case reporting to CPSP is conducted on a voluntary basis and not all SARS-CoV-2 hospitalizations were identified. Reporting fatigue towards the end of the study period may have biased case reporting in favour of more severe cases and therefore, severity outcomes may be overestimated. Moreover, testing indications may have varied by jurisdiction throughout the study period, such that not all admissions may have been tested for SARS-CoV-2 at all times. The reported minimum incidence proportions are therefore underestimates, but may still be useful to compare age-specific rates of hospitalization. Second, indications for ICU transfer may differ by centre and some children may have been admitted to ICU for precautionary purposes only. In this study, nine (of 98) children met criteria for severe disease because of their ICU admission and otherwise lacked features of severe COVID-19 (e.g. respiratory support requirements beyond low-flow oxygen or organ system involvement). Third, as the pandemic progressed, the case report form could not be amended to include variables such as SARS-CoV-2 lineage, length of hospital stay, and maternal immunization status. Fourth, our analysis of risk factors was conducted within hospitalized patients only, and therefore may differ from risk factors of severe disease among all children with COVID-19. Analyses of risk factors could not be conducted within age strata and for example, effects of obesity may not be generalizable to children <5 years of age.^37^ Finally, population group of the child was reported by physicians and not by families, and therefore these variables could not be included in multivariable models.

In this national prospective surveillance study, severe COVID-19 was detected across all age groups and in children with and without comorbid conditions. Neurologic and pulmonary conditions, as well as technology dependence requirements were associated with higher risk of severe COVID-19. These findings may be used to inform vaccination and booster campaigns, prioritize allocation of COVID-19 therapies, and guide family and policymaker decision-making as the pandemic continues.

## RESEARCH IN CONTEXT

### Evidence before this study

We searched Medline and PubMed for published materials and medRxiv for pre-print publications between January 1, 2020 and February 28, 2022. Search terms used included the following combination of words and did not include language limitations: “COVID-19” OR “SARS-CoV-2” OR “coronavirus”; “children” OR “pediatric” OR “adolescent” OR “youth”; “hospitalized” or “admitted”; “severe” OR “critical” OR “intensive care” OR “complications” OR “mortality” OR “death” OR “risk factors”. We identified numerous descriptive studies describing clinical characteristics and outcomes of hospitalized children and youth, including analyses of single or multi-centre studies and population-based databases. Fewer studies (n=10) included multivariable analysis of severe COVID-19 risk factors. Chronic comorbid conditions were commonly associated with severe outcomes, including diabetes, neurologic conditions, and respiratory conditions. Age was frequently associated with COVID-19 severity, though with conflicting results particularly regarding younger children (i.e. <1 year).

### Added value of this study

The Canadian Paediatric Surveillance Program collected detailed data on clinical presentations and chronic comorbid conditions among nearly 60% of all paediatric SARS-CoV-2 hospitalizations in Canada between March 2020–May 2021. A large available sample size (n=330 COVID-19-related hospitalizations) permitted multivariable analysis of specific comorbid conditions. We confirm neurologic and pulmonary conditions as risk factors for severe disease, including specifically epilepsy, bronchopulmonary dysplasia, and uncontrolled asthma. We also identify greater risk among children with technology dependence requirements, as well as children aged 2–4-years and 16–<18-years.

### Implications of all the available evidence

Children at all ages and both with and without comorbid conditions may experience severe COVID-19 outcomes. Conditions associated with medical complexity, including neurologic and pulmonary conditions as well as technology dependence, are at heightened risk of severe disease. Children <5 years remain a key unvaccinated group both in Canada and globally. These findings may help inform clinical practice, including the use of targeted COVID-19 therapies to reduce further morbidity and mortality.

## Data Availability

De-identified data that underlie the results reported in this article (text, tables, figures and appendices) and that abide by the privacy rules of the Canadian Paediatric Surveillance Program and the Public Health Agency of Canada can be made available to investigators whose secondary data analysis study protocol has been approved by an independent research ethics board.

## AUTHOR CONTRIBUTIONS

The study was conceived by DSF, OD, CMH, FK, and SKM. DSF conducted statistical analysis and wrote the first draft of the manuscript. All authors had access to the data, and DSF, CMH, MLT, MK, and SKM have accessed and verified the data underlying the study. All authors contributed to data collection, reviewed the study results and manuscript, and approved of the final manuscript.

## ACKNOWLEDGEMENTS

The authors wish to thank the paediatricians, paediatric subspecialists, and health professionals who voluntarily respond to CPSP surveys. The authors are also grateful to the staff and managers of the CPSP for their commitment to this study, as well as members of the CPSP Scientific Steering Committee who serve as stewards of the program. They also thank the members and leadership of the Paediatric Inpatient Research Network for cases reported and their dedication to the CPSP.

