## Supplementary materials for "Risk factors for severe COVID-19 in hospitalized children in Canada: A national prospective study from March 2020–May 2021"

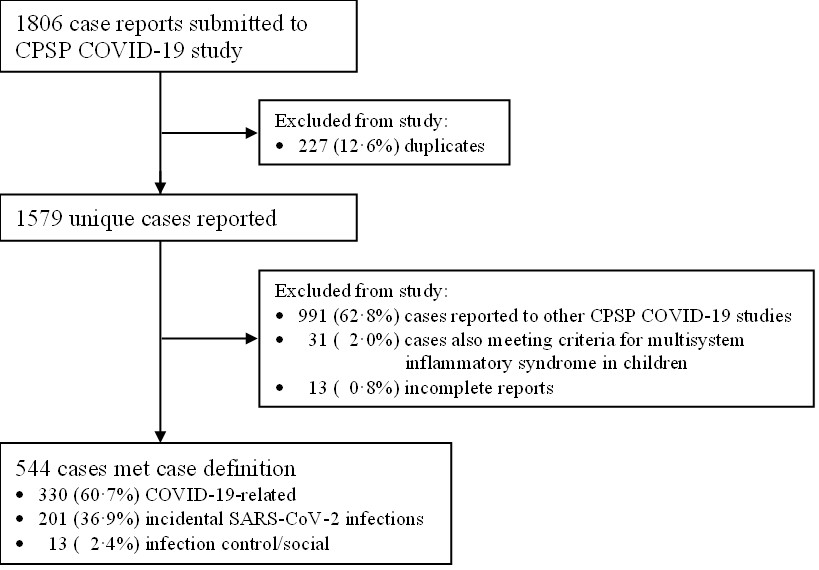
**Supplementary Figure S1.** Flow chart of cases reported to the CPSP COVID-19 study.

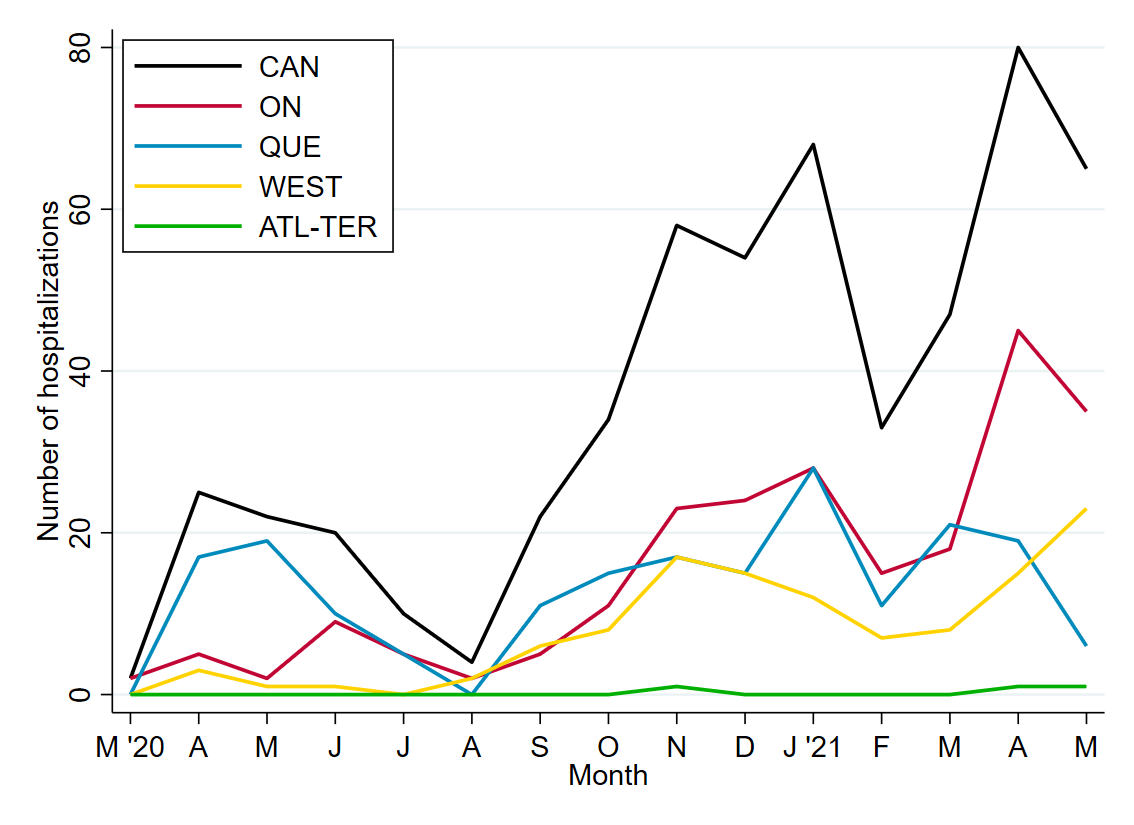

**Supplementary Figure S2. National and subnational <18-year SARS-CoV-2 hospitalizations by calendar month during CPSP study period.**

CAN=Canada (n=544); ON=Ontario (n=229); QUE=Quebec (n=194); WEST=Western Canada (n=118), including Alberta, British Columbia, Manitoba, and Saskatchewan; ATL-TER=Atlantic Canada and Territories (n<5), including New Brunswick, Newfoundland and Labrador, Northwest Territories, Nova Scotia, Nunavut, Prince Edward Island, and Yukon.

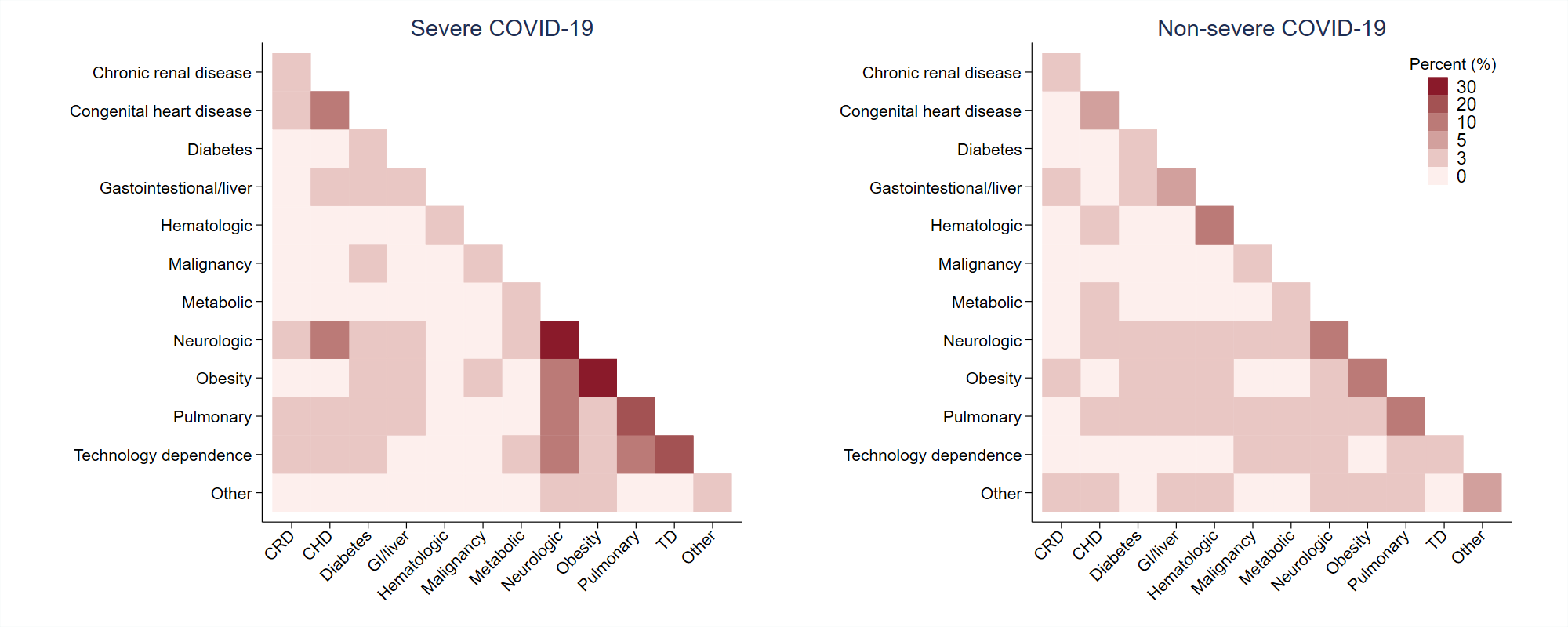
**Supplementary Figure S3. Overlap of chronic conditions in patients by COVID-19 disease severity.**

The diagonal represents the percentage of patients with each comorbidity category. CRD=Chronic renal disease; CHD=Congenital heart disease; TD=Technology dependence.

| **Supplementary Table S1. Chronic comorbid conditions among all SARS-CoV-2 hospitalizations.** | | | | |
| --- | --- | --- | --- | --- |
| **Chronic comorbid conditions, n (%)** | | Hospitalized due to COVID-19-related disease | | *P* value |
|  |  | Yes | No^1^ |  |
| **Number of hospitalizations, N** | | 330 | 214 | --- |
| **Neurologic/neurodevelopmental disorder, any^2^** | | 46 (13·9) | 39 (18·2) | 0·18 |
|  | Epilepsy | 20 (6·1) | 13 (6·1) | 0.99 |
|  | Chronic encephalopathy (e.g. cerebral palsy) | 19 (5·8) | 12 (5·6) | 0.94 |
|  | Chromosomal/genetic disorders | 9 (2·7) | 10 (4·7) | 0.23 |
|  | Autism spectrum disorder | 5 (1·5) | 5 (2·3) | 0.49 |
|  | Neurologic, NOS | 7 (2·1) | 9 (4·2) | 0.16 |
| **Obesity^3^** | | 44 (13·3) | 8 (3·7) | <0·001 |
| **Pulmonary, any^2^** | | 34 (10·3) | 7 (3·3) | 0·002 |
|  | Asthma | 16 (4·8) | <5 (<2·3) | 0.03 |
|  | Bronchopulmonary dysplasia | 9 (2·7) | <5 (<2·3) | 0.21 |
|  | Recurrent aspiration pneumonia | 5 (1·5) | <5 (<2·3) | 0.71 |
|  | Pulmonary, NOS | 5 (1·5) | <5 (<2·3) | 0.71 |
| **Immunosuppression, any^2,4^** | | 19 (5·8) | 16 (7·5) | 0·42 |
|  | Immunocompromising medications | 14 (4·2) | 14 (6·5) | 0.24 |
|  | Immunosuppression, NOS | 7 (2·1) | <5 (<2·3) | 0.75 |
| **Hematologic disease, any^2^** | | 17 (5·2) | 8 (3·7) | 0·44 |
|  | Sickle cell disease | 14 (4·2) | <5 (<2·3) | 0.13 |
|  | Hematologic, NOS | 5 (1·5) | <5 (<2·3) | 0.74 |
| **Technology dependence, any^2,5^** | | 16 (4·8) | 5 (2·3) | 0·14 |
|  | Parenteral nutrition | 12 (3·6) | <5 (<2·3) | 0.052 |
|  | Respiratory technology dependence | 7 (2·1) | <5 (<2·3) | 0.49 |
| **Congenital heart disease** | | 14 (4·2) | 7 (3·3) | 0·57 |
| **Gastrointestinal/liver disease** | | 10 (3·0) | 14 (6·5) | 0·051 |
| **Chronic renal disease** | | 9 (2·7) | <5 (<2·3) | 0·38 |
| **Diabetes mellitus** | | 8 (2·4) | 10 (4·7) | 0·15 |
| **Malignancy, any** | | 8 (2·4) | 9 (4·2) | 0·24 |
|  | Leukemia | 6 (1·8) | 6 (2·8) | 0.44 |
| **Metabolic disease** | | 8 (2·4) | <5 (<2·3) | 0·10 |
| **Bone diseases** | | <5 (<1·5) | <5 (<2·3) | 0·31 |
| **Other perinatal conditions** | | <5 (<1·5) | <5 (<2·3) | >0·99 |
| **Psychiatric disorder** | | <5 (<1·5) | 9 (4·2) | 0·001 |
| **Transplant recipient** | | <5 (<1·5) | <5 (<2·3) | 0·65 |
| NOS = Not otherwise specified. | | | | |
| ^1^Includes patients hospitalized with incidental infection or admitted for infection control or social purposes. | | | | |
| ^2^Multiple specific conditions could be reported (e.g. two or more neurologic conditions) and therefore may not sum to category total. | | | | |
| ^3^Obesity was physician-reported and could not be confirmed for children missing height and weight data (22 COVID-19-related and 4 non-COVID-19-related hospitalizations). | | | | |
| ^4^Defined as immunocompromising medications, primary or secondary immunodeficiency otherwise specified, or chronic rheumatologic or autoimmune disorder. | | | | |
| ^5^Defined as parenteral nutrition, respiratory technology requirements (i.e. home oxygen or tracheostomy), or dialysis. | | | | |

| **Supplementary Table S2. Chronic comorbid conditions by age group.** | | | | | | |
| --- | --- | --- | --- | --- | --- | --- |
| **Characteristics** | | Child age | | | | |
|  |  | <6 months | 6–23 months | 2–4 years | 5–11 years | 12–17 years |
| **COVID-19-related hospitalizations, N** | | 123 | 46 | 39 | 29 | 92 |
| **Any chronic conditions, n (%)** | |  |  |  |  |  |
|  | None/Unknown | 111 (90·2) | 23 (50·0) | 21 (53·9) | 7 (23·3) | 26 (28·3) |
|  | One or more | 12 (9·8) | 23 (50·0) | 18 (46·1) | 23 (76·7) | 66 (71·7) |
| **Chronic condition category, n (%)** | |  |  |  |  |  |
|  | None/Unknown | 111 (90·2) | 23 (50·0) | 21 (53·9) | 7 (23·3) | 26 (28·3) |
|  | Non-complex | 8–11 (6·5–8·9) | 8 (17·4) | 8 (20·5) | 11 (36·7) | 41 (44·6) |
|  | Complex | <5 (<4·1) | 15 (32·6) | 10 (25·6) | 12 (40·0) | 25 (27·2) |

| **Supplementary Table S3. Severity and treatment outcomes by pandemic wave.** | | | | | |
| --- | --- | --- | --- | --- | --- |
| **Characteristics** | | Pandemic wave | | | *P* value |
|  |  | First wave; Mar–Aug 2020 | Second wave; Sep 2020–Feb 2021 | Third wave; Mar–May 2021 |  |
| **COVID-19-related hospitalizations, N** | | 37 | 165 | 128 | --- |
| **Age (years)^1^, median (IQR)** | | 1·5 (0·1–12·8) | 1·6 (0·1–12·0) | 2·2 (0·3–14·5) | 0·17 |
| **COVID-19 severity, n (%)** | |  |  |  | 0·10 |
|  | Mild illness | 25 (67·6) | 97 (58·8) | 59 (46·1) |  |
|  | Moderate illness | 5 (13·5) | 24 (14·5) | 22 (17·2) |  |
|  | Severe illness | 7 (18·9) | 44 (26·7) | 47 (36·7) |  |
| **Admitted to ICU, n (%)** | | 6 (16·2) | 29 (17·6) | 25 (19·5) | 0·86 |
|  | Length of ICU stay, median (IQR) | 6 (1–7) | 4 (2–8) | 3 (2–6) | 0·95 |
| **Respiratory/hemodynamic support required, n (%)^2^** | | 8 (21·6) | 43 (26·1) | 57 (44·5) | 0·001 |
|  | Low-flow oxygen | 6 (16·2) | 21 (12·7) | 31 (24·2) |  |
|  | High-flow nasal cannula | <5 (<13·5) | 12–15 (7·3–9·1) | 16–19 (12·5–14·8) |  |
|  | Non-invasive ventilation (e.g. CPAP or BiPAP) | <5 (<13·5) | 5–8 (3·0–4·8) | 5–8 (3·9–6·3) |  |
|  | Conventional mechanical ventilation | <5 (<13·5) | 9–12 (5·5–7·3) | 9–12 (7·0–9·4) |  |
|  | Vasopressors | <5 (<13·5) | <5 (<3·0) | <5 (<3·9) |  |
|  | Extracorporeal membrane oxygenation | 0 (0·0) | 0 (0·0) | 0 (0·0) |  |
| **COVID-19-related therapies** | |  |  |  |  |
|  | Steroids | 5 (13·5) | 37 (22·4) | 46 (35·9) | 0·005 |
|  | Anticoagulation | <5 (<13·5) | 10–13 (6·1–7·9) | 22–25 (17·2–19·5) | 0·009 |
|  | Remdesivir | <5 (<13·5) | 10–13 (6·1–7·9) | 21–24 (16·4–18·8) | 0·004 |
| BiPAP=Bilevel positive airway pressure; CPAP=Continuous positive airway pressure; ICU=Intensive care unit; IQR=Interquartile range. | | | | | |
| ^1^Continuous age missing for 4 cases (1 second wave, 3 third wave). | | | | | |
| ^2^Multiple supports could be reported and therefore specific supports do not sum to any supports. | | | | | |

| **Supplementary Table S4. Comparison of modified WHO and modified Dong severity criteria, as applied in this study.** | | | | |
| --- | --- | --- | --- | --- |
| **Category** | | **Frequency (N)** | | **Percent (%)** |
| **COVID-19-related hospitalizations, N** | | 329 | | --- |
| **COVID-19 severity, modified WHO algorithm^1^** | | |  | |
|  | Mild disease | 181 | | 54·9 |
|  | Moderate disease | 51 | | 15·5 |
|  | Severe disease | 98 | | 29·7 |
| **COVID-19 severity, modified Dong algorithm^2^** | |  | |  |
|  | Mild disease | 96 | | 29·1 |
|  | Moderate disease | 50 | | 15·2 |
|  | Severe disease | 74 | | 22·4 |
|  | Critical disease | 110 | | 33·3 |
| ^1^Modified from the WHO Working Group on the Clinical Characterisation and Management of COVID-19 infection (2020). | | | | |
| ^2^Modified from Dong et al (2020). | | | | |

**APPENDIX 1**

| **Severity category** | **Criteria^1^** |
| --- | --- |
| **Mild disease** | Patient meets all of the following criteria:   - Section 4.2: Symptomatic COVID-19 including any of fever, cough, sore throat, runny nose, sneezing, lethargy, skin manifestations, muscle aches, rash, vomiting, diarrhea, loss of appetite, conjunctivitis, headache, loss of smell, or loss of taste. - Does not meet criteria for moderate or severe disease. |
| **Moderate disease** | Patient meets the following criteria:   - Section 4.2: Symptomatic COVID-19 including any of fever, cough, sore throat, runny nose, sneezing, lethargy, skin manifestations, muscle aches, rash, vomiting, diarrhea, loss of appetite, conjunctivitis, headache, loss of smell, or loss of taste.   AND one or both of the following:   - Section 7.1: Administered COVID-19 targeted therapy including any of remdesivir, corticosteroids, hydroxychloroquine, chloroquine, anti-IL-1, or anti-IL-6. - Section 7.4: Administered increased baseline home oxygen or low-flow oxygen.   AND   - Does not meet criteria for severe disease. |
| **Severe disease** | Patient meets any of the following:   - Section 4.2: Clinical features including coma or seizures. - Section 4.4: Clinical features including acute respiratory distress syndrome, cytokine storm/macrophage activating syndrome, seizures, stroke, encephalitis, encephalopathy, acute necrotizing encephalopathy, coma, hypotension, or acute cardiac dysfunction. - Section 7.3: Requires admission to intensive care unit. - Section 7.4: Administered high-flow nasal canula, non-invasive ventilation (e.g. CPAP or BiPAP), conventional mechanical ventilation, high-frequency oscillatory ventilation, nitric oxide, extracorporeal membrane oxygenation, or vasopressors. - Section 7.6: Death attributable to COVID-19. |

^1^Criteria adapted from World Health Organization^17^.
